# South to North Migration Patterns of Tuberculosis Patients Diagnosed in the Mexican Border with Texas

**DOI:** 10.1101/2021.03.31.21254658

**Authors:** Jennifer S. Curry, Bassent Abdelbary, Moncerrato García-Viveros, Juan Ignacio Garcia, Marcel Yotebieng, Adrian Rendon, Jordi B. Torrelles, Blanca I. Restrepo

## Abstract

**Background:** Immigration is a determinant of tuberculosis (TB) epidemiology. The US-Mexican border state of Tamaulipas serves as a migration waypoint for further immigration to the US, and has the second highest incidence of TB in Mexico. Here, we determined the contribution and characteristics of immigrants to the TB burden in Tamaulipas.

**Methods:** TB surveillance data from Tamaulipas (2006-2013) was used to conduct a cross-sectional characterization of TB immigrants (born outside Tamaulipas) and identify their association with TB treatment outcomes.

**Results:** Immigrants comprised 30.8% of the TB patients, with >99% originating from internal Mexican migration. Most migration was from South to North, with cities adjacent to the US border as destinations. Immigrants had higher odds of risk factors for TB [older age (≥ 65 yr old, OR 2.4, 95% CI 2.1, 2.8), low education (OR 1.3, 95% CI 1.2, 1.4), diabetes (OR 1.2, 95% CI 1.1, 1.4)], or abandoning TB treatment (adjusted OR 1.2, 95% CI 1.0, 1.5).

**Conclusions:** The US port of entry of Tamaulipas has a predominant south to north migration, positively impacting TB prevalence in this region. There is a need to identify strategies to prevent and manage TB more effectively in this Mexican migration waypoint.

## INTRODUCTION

Foreign-born individuals bear a high proportion of the overall tuberculosis (TB) burden in high-income countries^1^. In the US, immigrants account for 71.4% of all TB cases and 85% of the multidrug-resistant (MDR) TB cases, with Mexican nationals comprising nearly 20% in both cases^2,3^. The incidence of TB and MDR-TB in the US has significantly declined in the last decade, but this is not the case for the US states bordering Mexico^4-6^. And in the border state of Texas, the US counties bordering Mexico, Mexican nationals comprise more than 60% of the TB patients.^7^

The Mexican region bordering the US houses a high number of vulnerable populations including, drug users, illegal immigrants, and most recently, COVID-19 patients^8-12^. The border has historically attracted immigrants from TB endemic areas like Southern Mexico and Central America^6,13^. In recent years, immigrants have progressively accumulated in overcrowded confinements that favor *Mycobacterium tuberculosis* transmission. Not surprisingly, the six Mexican Border States (Baja California, Sonora, Chihuahua, Coahuila, Nuevo León, and Tamaulipas) account for 34.3% of all TB cases in Mexico, with TB incidence rates of up to 44.9/100,000 in 2016, compared to 13.8/100,000 nationally^14-17^.

Mexico is considered the most important migration corridor in the world, serving as a country of origin, a place of transit, and a destination for many immigrants expecting to enter the US^18,19^. Within Mexico, Tamaulipas is a highly dynamic state. Annually, more than 50,000 individuals emigrate from Tamaulipas to settle in other Mexican states, e.g. Nuevo León, Veracruz, San Luis Potosí and Coahuila, and approximately another 30,000 emigrate to the US^20,21^. Nonetheless, this population loss is quickly made up by a similar influx of approximately 100,000 immigrants coming into Tamaulipas each year in search of better job opportunities and improved or safer lifestyles, mostly from other Mexican states, where the economy is depressed such as Veracruz, San Luis Potosí and Chiapas^20^.

Among the Mexican-US border states, Tamaulipas has the second highest burden of TB, and because of its geographical location, it serves as a migration waypoint. Elucidation of the migration patterns of TB patients in Tamaulipas would provide a deeper understanding of their contribution to TB incidence in this border state, and percolation to the US, but such studies are missing. The primary objective of this study was to determine the contribution of immigrants to increased TB cases in Tamaulipas, and identify their place of origin, migration destinations, sociodemographic characteristics and their likelihood of TB treatment success, when compared to local residents.

## MATERIAL AND METHODS

### Ethical committee

This study was approved by the institutional review board of the University of Texas Health Science Center at Houston (Human Subject IRB # HSC-SPH-15-0489) and the Research Committee for the Secretaría de Salud de Tamaulipas (Human Subjects IRB# 076/2015/CEI). This study followed the Strengthening the Reporting of Observational Studies in Epidemiology (STROBE) reporting guidelines.

### Data source and study population

We used data collected for TB surveillance purposes between 2006 and 2013 by the Mexican Health Department (Secretaría de Salud de Mexico) for the state of Tamaulipas. These data included all the new TB cases among patients 18 years and older. Definitions were based on the official Mexican National TB program. Namely, TB diagnosis was based on the detection of acid-fast bacilli by smear microscopy or histopathology, culture or clinical findings. Further details on the procedure for selection of 8,431 adults (18 years or older) with newly diagnosed TB patients were reported elsewhere ^22^.

### Measures and definitions

The primary exposure of interest was immigrant status of the TB patient, defined as “YES” for individuals reporting a place of birth outside of Tamaulipas. The dataset options included each Mexican state or “other countries”. Adverse treatment outcomes included failed TB treatment (when the patient remains smear positive after five months of anti-TB drug treatment), relapse (when presenting new TB within one year of treatment completion), abandon (when lost to follow-up for more than three months), or death (during the course of TB treatment). Treatment was evaluated by merging individual outcomes and categorizing them as successful or not per World Health Organization (WHO) definitions.^23^ Drug-resistant (DR) TB refers to resistance to any first-line antibiotic for TB, and MDR-TB indicates resistance to both isoniazid and rifampicin. Patient’s characteristics, previously described in detail^22^, included age, sex, occupation, undernutrition (determined clinically), diabetes (self-reported or blood confirmed), and alcoholism (self-reported). Tamaulipas is divided into different sanitary jurisdictions, with those adjacent to the US referred to as “border” and others as “non-border” (**Table S1**).

### Statistical analysis

We used proportions or means to describe the distribution of variables. We first evaluated three aspects of migration among TB patients in Tamaulipas: The proportion of immigrant TB patients, their place of origin, and their predominant migration routes. The Chi-square test for trend was used to detect TB incidence rate changes over the eight-year period of study. The percent change over time was estimated using both the linear interpolation method and average annual percent change to account for year-to-year change. Associations between immigrant status and their sociodemographics, comorbidities and TB treatment outcomes were also evaluated. Univariable logistic regression models were employed to estimate odds ratios (OR) and 95% confidence intervals assessing the differences in outcome across levels of co-variables. All multivariable logistic regression analysis included age and sex, and additional analysis was conducted with education status and comorbidities, in order to identify the independent contribution of place of birth to TB treatment success adjusted for potential confounders. All statistical tests were performed using STATA 14 (StataCorp. 2015. Stata Statistical Software: Release 14. College Station, TX: StataCorp LP) at a 5% level of significance.

## RESULTS

### Participants characteristics

The characteristics of the study population were described previously.^22^ Essentially, the mean age of the 8,431 TB patients was 43 years and 65.5% were males. Nearly half were unemployed and 58% had at most a primary school education. More than 90% had pulmonary TB and 15.2% had an adverse TB treatment outcome. One fourth had diabetes and 5.3% had HIV.

We found that the average proportion of immigrants with TB was 30.8% over the eight years studied (range 18.6% - 37.8%; **Table S1**). There were minor changes in the proportion of immigrants with TB over time, with the lowest in 2006 (18.6%) and 2007 (26.4%), and higher values and high between 2008 and 2013 [(range 30.8 – 37.8%); trend *p* value 0.526]. The TB incidence background was maintained similar over the eight-year study period (range 30.0 – 33.8/100,000 per year; trend *p* value 0.900; **Table S1**).

### Immigration patterns

Nearly all TB immigrants were born in other Mexican states (2,577/2,596, 99.3%) with less than 1% originating from other countries. Nearly three-quarters (1,906 of 2,596, 73.4%) were residing in border jurisdictions at the time of their TB diagnosis, with most in the cities of Reynosa (29.3%), Nuevo Laredo (22.3%) and Matamoros (16.8%; **Figure 1)**. Most immigrant were born in four Mexican states: Chiapas (n=181, 2.2%), Nuevo Leon (n=255, 3.0%), San Luis Potosi (n=476, 5.7%), and Veracruz (n=903, 10.7%). The most frequent destinations for immigrants originating from these four cities were as follows: TB patients originating from Chiapas were now mostly living in Nuevo Laredo (107/181, 59.1%), Reynosa (34/181, 18.8%), and Matamoros (23/181, 12.7%). Those from Nuevo Leon were now mostly in Reynosa (85/255, 33.3%) and Nuevo Laredo (83/255, 32.5%). Those from San Luis Potosi had mainly emigrated to Matamoros (124/476, 26.1%), Tampico (113/476, 23.7%), Nuevo Laredo (83/476, 17.4%), and Reynosa (70/476, 14.7%). And the TB patients born in Veracruz were mainly residing in Reynosa (367/903, 40.6%), Tampico (208/903, 23%), Matamoros (129/903, 14.3%), and Nuevo Laredo (83/903, 9.2%) (**Figure 1**).

**Figure 1.**
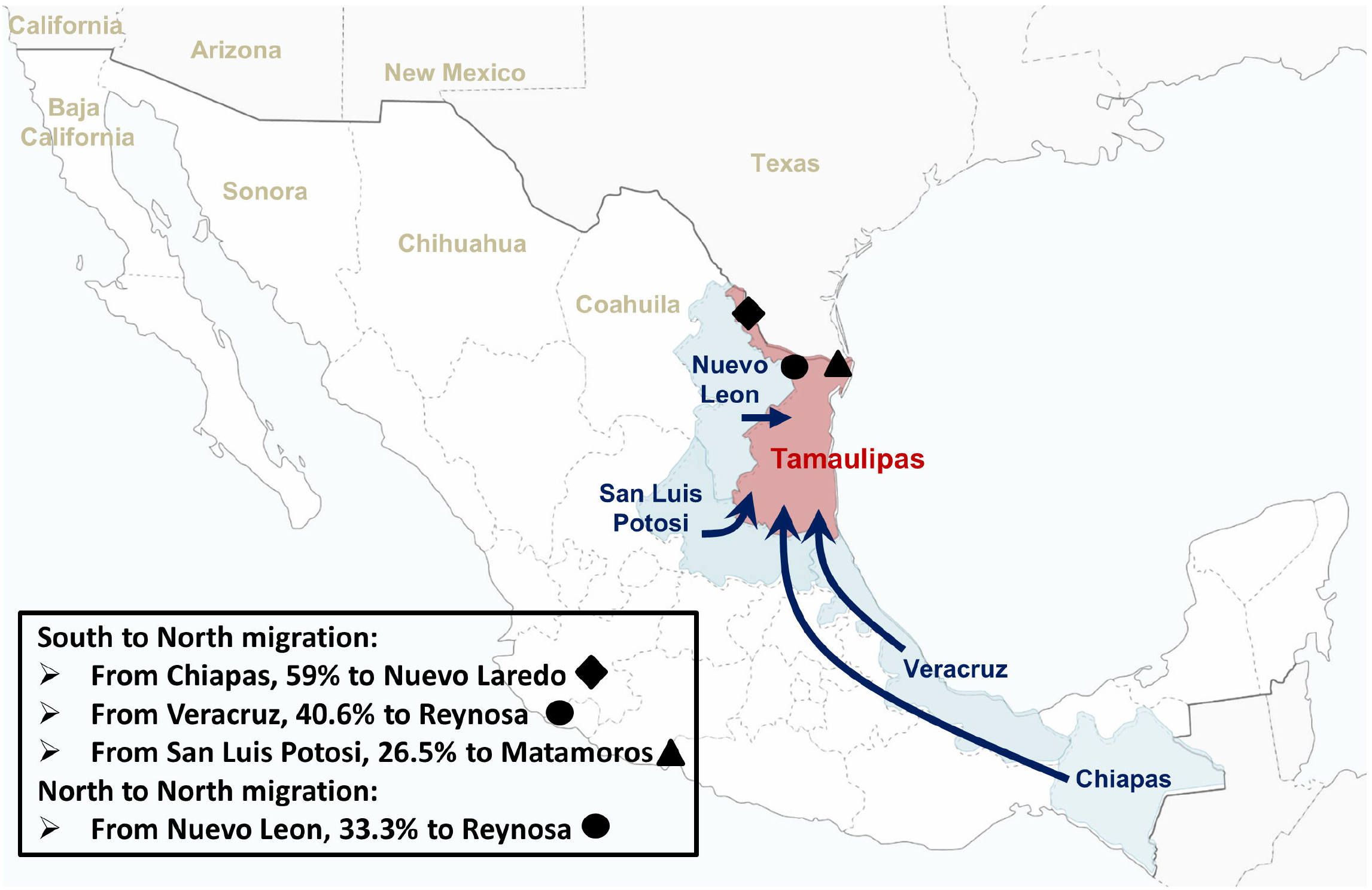
Predominant south to north migration patterns of TB patients from Tamaulipas. The predominant migration patterns of TB patients diagnosed in Tamaulipas is from South and North, and to a lesser extent, from North to North. The south to north routes were from Chiapas (in the Southern border with Guatemala) to Nuevo Laredo, or Veracruz to Reynosa or San Luis Potosi to Matamoros. The north to north pattern was from Nuevo Leon to Reynosa. In all cases, the final destination cities, namely Matamoros, Reynosa and Nuevo Laredo, were in northern Tamaulipas and adjacent to the US border. The proportion of immigrants from each place of origin, towards the indicated destination, is shown in the legend for each pathway. The names of the other states along the US-Mexico border as shown as a reference in lighter font.

### Characteristics of TB patients associated with immigrant status by place of birth

Compared to local residents, immigrant TB patients born outside of Tamaulipas were older [(OR 1.5, 95% CI 2.3, 1.6 for 41-64 yr old *vs*. OR 2.4, 95% CI 2.1, 2.8 for those older than 64 years], had a lower level of education (OR 1.3, 95% CI 1.2, 1.4, up to primary school *vs*. higher education), decreased odds of BCG vaccination (OR 0.9, 95% CI 0.8, 0.98), and/or decreased odds of having a positive smear for acid-fast bacilli at the time of TB diagnosis (OR 0.8, 95% CI 0.7, 0.9; **Table 1**). When evaluating comorbidities, immigrants with TB *vs*. those born in Tamaulipas had increased odds of diabetes (OR 1.2, 95% CI 1.1, 1.4), and lower HIV/AIDS (OR 0.7, 95% CI 0.6, 0.9). When adjusting for sociodemographics, a lower odds of a positive AFB smear or higher odds of abandoning treatment remained significant (**Table 1**).

**Table 1:** Demographic and clinical characteristics of TB patients by place of birth in Tamaulipas, 2006-2013 ^1^

| Characteristics | Born in Tamaulipas<br>(n = 5,835) | Not born in Tamaulipas<br>(n = 2,596) | OR (95% CI)<sup>2</sup> | Adj-OR (95% CI)<sup>3</sup> |
| --- | --- | --- | --- | --- |
| **Sociodemographics and comorbidities** |  |  |  |  |
| **Age** |  |  |  |  |
| 18 - 40 years old | 3,038 (52.1%) | 1,023 (39.4%) | 1.0 |  |
| 41 - 64 years old | 2,219 (38%) | 1,108 (42.7%) | **1.5 (1.3, 1.6)** |  |
| ≥ 65 years old | 578 (9.9%) | 465 (17.9%) | **2.4 (2.1, 2.8)** |  |
| **Male sex** | 3,849 (66%) | 1,674 (64.5%) | 0.9 (0.8, 1.03) |  |
| **Education** |  |  |  |  |
| No or primary education | 3,103 (55.9%) | 1,574 (62.1%) | **1.3 (1.2, 1.4)** |  |
| Higher education | 2,447 (44.1%) | 960 (37.9%) | 1.0 |  |
| **Occupation** |  |  |  |  |
| Employed | 1,848 (42.9%) | 781 (41.2%) | 1.0 | 1.0 |
| Not employed | 1,788 (41.5%) | 843 (44.5%) | 1.1 (0.9, 1.3) | 0.9 (0.8, 1.0) |
| Unemployed | 669 (15.5%) | 271 (14.3%) | 0.9 (0.8, 1.1) | 0.9 (0.7, 1.0) |
| **BCG (Yes)** | 4,211 (77%) | 1,783 (74.6%) | **0.9 (0.8, 0.98)** | 1.1 (0.9, 1.2) |
| **Comorbidities** |  |  |  |  |
| Diabetes | 1,397 (23.9%) | 724 (27.9%) | **1.2 (1.1, 1.4)** | 01.0 (0.9, 1.1) |
| HIV/AIDS | 332 (5.7%) | 113 (4.3%) | **0.7 (0.6, 0.9)** | 0.9 (0.7, 1.1) |
| Excess Alcohol use | 344 (5.9%) | 130 (5%) | 0.8 (0.7, 1.1) | 0.8 (0.7, 1.0) |
| Undernutrition | 474 (8.1%) | 226 (8.7%) | 1.1 (0.9, 1.3) | 1.1 (0.9, 1.3) |
| **Clinical presentation of TB** |  |  |  |  |
| **Location of disease** |  |  |  |  |
| Extra-pulmonary | 470 (8.1%) | 206 (7.9%) | 1.0 | 1.0 |
| Pulmonary | 5,364 (91.9%) | 2,390 (92.1%) | 1.0 (0.8, 1.2) | 0.9 (0.8, 1.2) |
| **AFB smear** | 4,629 (85.8%) | 1,982 (83.5%) | **0.8 (0.7, 0.9)** | **0.8 (0.7, 0.9)** |
| **Any drug resistance** | 179 (3.1%) | 80 (3.1%) | 1.0 (0.8, 1.3) | 1.0 (0.8, 1.4) |
| **Multi-drug resistance** | 40 (0.7%) | 15 (0.6%) | 0.8 (0.5, 1.5) | 0.9 (0.5, 1.7) |
| **Treatment outcomes** |  |  |  |  |
| Cured | 4,874 (85.1%) | 2,135 (83.9%) | 1.0 | 1.0 |
| Treatment failure | 107 (1.9%) | 51 (2%) | 1.1 (0.8, 1.5) | 1.1 (0.7, 1.5) |
| Death | 354 (6.2%) | 167 (6.6%) | 1.1 (0.9, 1.3) | 0.9 (0.7, 1.1) |
| Abandon treatment | 390 (6.8%) | 191 (7.5%) | 1.1 (0.9, 1.3) | **1.2 (1.0, 1.5)** |
| TB relapse | 126 (2.2%) | 48 (1.9%) | 0.8 (0.6, 1.2) | 0.8 (0.6, 1.2) |

<sup>1</sup> Data expressed as n (column %), with % based on the total number of cases with available information;
<sup>2</sup> Tamaulipas is the reference category; <sup>3</sup> Adjusted for age, sex, and education level; Bold p-values are significant;

Analysis was also conducted based on the most frequent states of birth: Chiapas, Nuevo Leon, San Luis Potosi or Veracruz (**Table 2**). We found that the odds of HIV/AIDS was lower in immigrants from Chiapas (OR 0.2, 95% CI 0.04, 0.7) or San Luis Potosi (OR 0.5, 95% CI 0.3, 0.9). Type 2 diabetes was more prevalent in those born in San Luis Potosi (OR 1.6, 95% CI 1.3, 1.9) and Veracruz (OR 1.2, 95% CI 1.1, 1.4). Regarding TB clinical characteristics, there were no differences between local and immigrant TB patients in the prevalence of the pulmonary location, drug resistant TB or smear results, except for lower smear positivity for immigrants with TB from Veracruz. Regarding TB treatment outcomes, when compared to TB patients born in Tamaulipas (locals), immigrant born in Nuevo Leon had a 2-fold increased odds of TB treatment failure, and those born in Chiapas had 2.1-fold increased odds of TB relapse. When controlling for age, sex, education level, BCG vaccination, diabetes and HIV/AIDS, being an immigrant from Nuevo Leon remained associated with abandonment of treatment (OR 1.9, 95% CI 1.2, 2.9; **Table 1**).

**Table 2.** Association between the most common places of birth of migrants, and sociodemographics, comorbidities, TB presentation and treatment outcomes ^1, 2^

| Place of birth | Chiapas<br>(n = 181) | Nuevo Leon<br>(n = 255) | San Luis Potosi<br>(n = 476) | Veracruz<br>(n = 903) |
| --- | --- | --- | --- | --- |
| **Sociodemographics and comorbidities** |  |  |  |  |
| **Age** |  |  |  |  |
| 18 - 40 years old | 1.0 | 1.0 | 1.0 | 1.0 |
| 41 - 64 years old | **1.5 (1.1, 2.2)** | **2.1 (1.6, 2.8)** | **2.3 (1.8, 2.8)** | 1 (0.9, 1.2) |
| ≥ 65 years old | **3.5 (2.3, 5.2)** | **3.6 (2.5, 5.1)** | **4.4 (3.4, 5.7)** | 0.9 (0.8, 1.3) |
| **Male sex** | 1.2 (0.9, 1.6) | 0.9 (0.7, 1.1) | 0.9 (0.7, 1.1) | 0.9 (0.8, 1.0) |
| **Education** |  |  |  |  |
| Higher education | 1.0 | 1.0 | 1.0 | 1.0 |
| Up to 1o education | **1.8 (1.3, 2.5)** | **1.6 (1.2, 2.0)** | **1.9 (1.5, 2.3)** | 0.9 (0.8, 1.1) |
| **Occupation** |  |  |  |  |
| Employed | 1.0 | 1.0 | 1.0 | 1.0 |
| Not employed | 1.4 (0.9, 2.0) | 1.3 (0.9, 1.8) | 1.2 (0.9, 1.6) | 0.9 (0.8, 1.1) |
| Unemployed | 1.2 (0.7, 2.0) | 1.2 (0.7, 1.8) | 1.2 (0.8, 1.6) | 0.9 (0.7, 1.1) |
| **BCG (Yes)** | 0.8 (0.6, 1.2) | 1 (0.8, 1.4) | 1.1 (0.8, 1.3) | 0.9 (0.8, 1.1) |
| **Comorbidities** |  |  |  |  |
| Diabetes | 0.9 (0.6, 1.3) | 1.2 (0.9, 1.6) | **1.6 (1.3, 1.9)** | **1.2 (1.1, 1.4)** |
| HIV | **0.2 (0.04, 0.7)** | 0.5 (0.3, 1.1) | **0.5 (0.3, 0.9)** | 1.3 (0.9, 1.7) |
| Excess Alcohol use | 0.7 (0.4, 1.5) | 0.8 (0.5, 1.5) | 0.9 (0.6, 1.4) | 0.8 (0.6, 1.1) |
| Malnutrition | 1.3 (0.8, 2.1) | 0.8 (0.5, 1.3) | 1 (0.7, 1.4) | 1.1 (0.9, 1.4) |
| **Clinical presentation of TB** |  |  |  |  |
| **Location of disease** |  |  |  |  |
| Extra-pulmonary | 1.0 | 1.0 | 1.0 | 1.0 |
| Pulmonary | 1.5 (0.8, 2.9) | 1.2 (0.7, 1.9) | 1.1 (0.8, 1.6) | 0.9 (0.7, 1.2) |
| **Diagnostic AFB smear** | 1.2 (0.7, 1.9) | 0.9 (0.6, 1.3) | 0.8 (0.6, 1.1) | **0.7 (0.6, 0.8)** |
| **Any drug resistance** | 0.7 (0.3, 1.9) | 1.2 (0.6, 2.3) | 0.6 (0.3, 1.2) | 0.8 (0.5, 1.2) |
| **Multi-drug resistance** | 1.6 (0.4, 6.7) | 1.1 (0.3, 4.8) | 0.9 (0.3, 2.9) | 0.3 (0.1, 1.3) |
| **Treatment outcomes** |  |  |  |  |
| Cured | 1.0 | 1.0 | 1.0 | 1.0 |
| Treatment failure | 0.9 (0.3, 2.9) | **2.0 (1.1, 4.1)** | 1.3 (0.7, 2.5) | 0.8 (0.4, 1.4) |
| Death | 1.2 (0.7, 2.1) | 1.0 (0.6, 1.8) | 1.1 (0.8, 1.6) | 0.9 (0.7, 1.3) |
| Abandon treatment | 1.1 (0.6, 1.9) | 1.5 (0.9, 2.3) | 0.6 (0.4, 1.0) | 1.2 (0.9, 1.6) |
| TB relapse | **2.1 (1.1, 4.3)** | 1.3 (0.6, 2.8) | 0.7 (0.3, 1.5) | **0.5 (0.3, 0.9)** |

<sup>1</sup> Tamaulipas is the reference category; <sup>2</sup> Data expressed as crude odds ratio (95% CI)

**Table 3.** Association between the place of birth of migrants and the clinical presentation and outcomes of TB ^1, 2^

| Place of birth | Chiapas<br>(n = 181) | Nuevo Leon<br>(n = 255) | San Luis Potosi<br>(n = 476) | Veracruz<br>(n = 903) |
| --- | --- | --- | --- | --- |
| **Location of disease** |  |  |  |  |
| Extra-pulmonary | 1.0 | 1.0 | 1.0 | 1.0 |
| Pulmonary | 1.3 (0.7, 2.7) | 1.1 (0.6, 1.8) | 0.9 (0.7, 1.4) | 0.9 (0.7, 1.3) |
| **Diagnostic AFB smear** | 1.1 (0.7, 1.8) | 0.8, (0.6, 1.2) | **0.7 (0.5, 0.9)** | **0.7 (0.6, 0.9)** |
| **Any drug resistance** | 0.6 (0.2, 2.0) | 1.1 (0.6, 2.3) | 0.7 (0.3, 1.4) | 0.8 (0.5, 1.2) |
| **Multi-drug resistance** | 1.2 (0.2, 8.8) | 0.93 (0.1, 5.1) | 1.0 (0.3, 3.5) | 0.3 (0.1, 1.6) |
| **Treatment outcomes** |  |  |  |  |
| Cured | 1.0 | 1.0 | 1.0 | 1.0 |
| Treatment failure | 1.1 (0.3, 3.4) | 2.0 (0.9, 4.1) | 1.2 (0.6, 2.4) | 0.9 (0.5, 1.7) |
| Death | 1.2 (0.6, 2.2) | 0.8 (0.4, 1.5) | 0.9 (0.6, 1.3) | 0.9 (0.7, 1.3) |
| Abandon treatment | 1.3 (0.7, 2.4) | **1.9 (1.2, 2.9)** | 0.7 (0.4, 1.2) | 1.2 (0.9, 1.6) |
| TB relapse | 1.4 (0.6, 3.5) | 1.3 (0.6, 2.9) | 0.6 (0.3, 1.4) | **0.5 (0.3, 1.0)** |

<sup>1</sup> Tamaulipas is the reference category; <sup>2</sup> Data expressed as odds ratio (95% CI), adjusted for for age, sex, education, BCG vaccination, diabetes and HIV/AIDS; AFB, Acid-fast bacilli

## DISCUSSION

We conducted a study aimed at understanding the epidemiology of TB among immigrants in Tamaulipas, a Mexican state bordering the US. We found that immigrants represented one-third of the TB patients, with >99% originating from other Mexican states. TB immigrants were older than local residents. This was unexpected and it is possible that they arrived when they were younger and now have a reactivation TB or a recent infection- we cannot distinguish both possibilities. Consistent with their older age, immigrants with TB had lower levels of education, higher prevalence of diabetes and lower BCG vaccination. TB immigrants worldwide are less likely to have treatment success, due in part to their higher likelihood of treatment interruption^24^. After adjustment, abandonment of treatment was higher.

Mexico has a total population of approximately 126 million, with close to 50% living in poverty^25,26^. Economic insecurity and drug and human trafficking cartels are main propagators of Mexican internal migration movements, with about 20% living in states in which they were not born ^26-29^. It is alluded that migration from the poorest regions in Mexico may be responsible for a significant proportion of TB cases in the Mexican states located across the US border, but this has never been systemically evaluated. A study conducted 25 years ago among immigrant TB patients identified at local health departments at the four US border states, provides some insights ^30^. Of the 164 Hispanic immigrant TB patients, 154 were born in Mexico and ten in Central America. Among the 154 born in Mexico, 93 (60%) had been living in a border Mexican town before emigrating to the US, and of these 93, 51 (55%) had first moved to a Mexican border town for at least 2 years before emigrating to the US. The findings from this study illustrates the influx of immigrants from non-border Mexican regions, into the border, and then into the USA, suggesting the importance of TB control in border and non-border regions of Mexico for control of TB in communities along the Texas-Mexico border ^30^.

A limitation of our study is that the analysis covered a period dating seven to fourteen years back. We do not have data on the proportion of immigrant TB patients in more recent years, but we would anticipate an increase. This is based on the regional trend for the increase in caravans of immigrants departing from South and Central American countries where TB is also endemic, with hopes of reaching the US, but with many remaining stuck in northern Mexico^31,32^. Tamaulipas is a “Remain in Mexico” (RIM) state under the Immigrant Protection Protocols (MPP) implemented in 2019 with top immigrant nationalities from Honduras, Guatemala, Cuba, El Salvador, and Ecuador. These policies highlight the epidemiological landscape of the Mexican border communities. Thus, we expect that in current times the contribution of immigrants to TB control would be at least the same or higher than our observations.

Another study limitation is that we conducted a secondary analysis of data collected for TB surveillance. Therefore, we were not able to calculate the TB incidence among those born in Tamaulipas versus immigrants born elsewhere. Year of immigration was not available either, so we could distinguish between migrants who were temporarily located in Tamaulipas *vs* immigrants who had permanently settled in this state. Furthermore, in our analysis we could not adjust for the years of residence prior to TB diagnosis. We did not have information on the reasons motivating migration, pre-departure health status, or travel conditions that can increase TB risk^33^. We cannot rule out reporting bias due to the stigma associated with migration. An association between immigrant status and drug-resistant TB was not measured. Another limitation was lack of access to the identification and treatment of latent TB infection, a key action in the pillar 1 of the current WHO End TB Strategy^34,35^.

In contemporary times an additional challenge to TB control is the coronavirus disease 2019 (COVID-19) pandemic. In Mexican border states, poverty, overcrowding, stress from immigration and lack of access to healthcare have contributed to high COVID-19 case-fatality rates in border Mexican states (16% compared to 8.5% nationally in 2020).^36^ Not surprisingly, COVID-19 has had a notable impact on TB services in the US-Mexico border. This, together with immigration patterns from South to North, is likely to result in higher drug susceptible and drug resistant TB cases in the upcoming years, with potential spill over to US border states.

In summary, we find that approximately one-third of the TB patients in Tamaulipas are immigrants from within Mexico, mostly originating from southern states and moving to communities along the US border. Given the recent increase in migration within Mexico due to drug-related cartels, or from Latin American countries ^37^, further studies are necessary to understand in more depth the underlying reasons, the detailed migration pathways, the prevalence of drug-resistance and the TB treatment outcomes in this mobile population. Understanding these issues will be essential to design strategies for improved TB control in gateway states like Tamaulipas. Such studies will also serve as a model to address the challenge of migration on TB control in other regions of the US-Mexico border, in the adjacent US where immigrants comprise a significant proportion of the TB patients, or in other countries worldwide where millions migrate each year from TB-endemic regions.^38^

## Data Availability

Data will be provided as per request

## Conflict of Interest

All authors declare no conflict of interest.

## Acknowledgments

We thank all the Tamaulipas state health jurisdictions who contributed to the collection and recording of TB data, Dr. Santa Elizabeth Ceballos Liceaga from the Sistema de Vigilancia Epidemiológica de Tuberculosis y Lepra, Dirección General de Epidemiología, Secretaría de Salud de México, for providing the surveillance datasets for our analysis.

## Funding

This research did not receive funding support from an agency, commercial or not-for-profit sectors. BIR has current funding support from NIH, NIA P01-AG051428 (PI Dr. Joanne Turner) and NIAID 1R21AI144541

## Authors contributions

**Jennifer S. Curry**: Conceptualization, Data analysis, interpretation, original draft of manuscript; **Bassent Abdelbary**: Data analysis, interpretation, edition of manuscript; **Moncerrato García-Viveros**: Conceptualization and coordination of data collection; **Juan I. García**: Data analysis interpretation and edition of manuscript; **Marcel Yotebieng**: Data analysis interpretation and edition of manuscript; **Adrian Rendon**: Data analysis interpretation and edition of manuscript; **Jordi B. Torrelles**: Data analysis interpretation and edition of manuscript; **Blanca I. Restrepo**: Conceptualization, Project administration, Resources, data interpretation, edition of manuscript. **All authors**: Reviewed and approved the final version of this manuscript.

## Notes

### Competing Interest Statement

The authors have declared no competing interest.

### Author Declarations

This study was approved by the institutional review board of the University of Texas Health Science Center at Houston (Human Subject IRB # HSC-SPH-15-0489) and the Research Committee for the Secretaria de Salud de Tamaulipas (Human Subjects IRB# 076/2015/CEI).

