## Supplementary material for "South to North Migration Patterns of Tuberculosis Patients Diagnosed in the Mexican Border with Texas": Supplmental tables S1 and Figure S1

**Classification of Tamaulipas health department into border and non-border regions**. Tamaulipas is sub-divided into twelve health districts referred to as sanitary jurisdictions that were categorized as “border” and “non-border” with respect to Texas. The non-border sanitary jurisdictions are Victoria, Tampico, Ciudad Mante, San Fernando, Jaumave, Padilla, and Altamira. The border included Matamoros, Reynosa, Nuevo Laredo, Miguel Aleman, and Valle Hermoso (**Table S1**).

**The TB incidence in Tamaulipas was unchanged over the study period**. We conducted a sensitivity analysis to examine if the annual TB incidence of Tamaulipas or within its sanitary jurisdictions had changed over time, and hence, would be associated with yearly migration proportions. To calculate annual TB incidence rates, total population of Tamaulipas or its sanitary jurisdictions was obtained using 2010 data from the Mexican Instituto Nacional de Estadística, Geografía e Informática (INEGI) report ^21^. Census data is updated every five years and 2010 was the closest midpoint to our study period. The Chi-square test for trend was used to detect TB incidence rate changes over the eight-year period of study.

We found that the TB incidence in Tamaulipas ranged between 30 and 33.8 per 100,000 per year during the 2008-2013 study period (**Table S1** and **Figure S1**). The border jurisdictions had higher TB incidence rates when compared to non-border regions (range 37.6 to 41.5 in border; 21.4 to 28.2 per 100,000 per year for non-border). The highest annual TB incidence rates were in Nuevo Laredo for the border and Tampico in the non-border regions. A Chi-square test for trend revealed no significant changes (either increase or decrease) in the entire state or in its border or non-border regions or most of its sanitary jurisdictions. The only exceptions were two jurisdictions located in the non-border region; Tampico and Altamira. For Tampico the trend analysis revealed an overall average annual borderline significant decrease of 4.5% (*p* = 0.053). Conversely, Altamira had a significant overall increase of 6.4% with an average annual percent change of 7.7% (*p* =0.027 **Table S1**).

| **Table S1. Proportion of migrant TB patients and TB incidence in Tamaulipas or its sanitary jurisdictions, by year** | | | | | | | | | |
| --- | --- | --- | --- | --- | --- | --- | --- | --- | --- |
|  | **2006** | **2007** | **2008** | **2009** | **2010** | **2011** | **2012** | **2013** | **Trend**  ***p*-value** |
| **Proportion of migrant TB patients** | | | | | | | | | |
| Tamaulipas | 18.6% | 26.4% | 37.8% | 33.4% | 34.6% | 33.4% | 30.8% | 31.8% | 0.526 |
| **TB incidence in Border jurisdictions ^1^** | | | | | | | | | |
| Matamoros | 36.4 | 34.1 | 37.0 | 36.0 | 35.8 | 39.5 | 34.1 | 37.0 | 0.655 |
| Reynosa | 42.0 | 40.7 | 42.0 | 37.1 | 47.6 | 37.4 | 46.3 | 36.8 | 0.568 |
| Nuevo Laredo | 44.0 | 47.9 | 43.2 | 47.9 | 48.2 | 56.5 | 42.2 | 54.2 | 0.342 |
| Miguel Aleman | 29.9 | 20.9 | 25.4 | 20.9 | 19.4 | 32.9 | 34.3 | 34.3 | 0.134 |
| Valle Hermoso | 27.0 | 27.0 | 36.4 | 28.1 | 30.3 | 29.8 | 30.3 | 27.6 | 0.355 |
| **TB incidence in non-border jurisdictions ^1^** | | | | | | | | | |
| Ciudad Victoria | 25.0 | 21.2 | 19.6 | 21.4 | 22.4 | 24.5 | 18.9 | 22.7 | 0.801 |
| Tampico | 40.4 | 33.1 | 32.3 | 30.1 | 37.0 | 31.7 | 31.7 | 27.7 | **0.053** |
| Ciudad Mante | 28.4 | 20.3 | 19.7 | 17.9 | 14.5 | 32.4 | 22.0 | 23.7 | 0.705 |
| San Fernando | 17.6 | 20.5 | 14.6 | 13.2 | 13.2 | 14.6 | 17.6 | 17.6 | 0.744 |
| Jaumave | 21.6 | 9.0 | 9.0 | 10.8 | 23.4 | 12.6 | 14.4 | 19.8 | 0.466 |
| Padilla | 27.3 | 15.8 | 12.9 | 18.7 | 15.8 | 11.5 | 15.8 | 11.5 | 0.091 |
| Altamira | 15.1 | 13.0 | 16.5 | 13.0 | 18.3 | 19.3 | 24.6 | 21.8 | **0.027** |
| **TB incidence totals  ^1^** | | | | | | | | | |
| Border jurisdictions | 38.8 | 38.3 | 39.6 | 37.6 | 41.5 | 41.3 | 39.8 | 39.6 | 0.154 |
| Non-border jurisdictions | 28.2 | 22.7 | 22.2 | 21.4 | 24.8 | 25.3 | 24.1 | 23.4 | 0.950 |
| Tamaulipas | 33.8 | 30.9 | 31.5 | 30.0 | 33.6 | 33.7 | 32.4 | 32.0 | 0.900 |
| ^1^ TB incidence per 100,000 per year with population size based on the report from the Instituto Nacional de Estadística, Geografía e Informática, INEGI; Bold p-values are significant | | | | | | | | | |

**Figure S1. Annual incidence of TB in Tamaulipas and by border regions. A.** TB Incidence in border and non-border sanitary jurisdications and in the entire state of Tamaulipas. **B.** TB incidence in border jurisdictions (Matamoros, Reynosa, Nuevo Laredo, Miguel Aleman, Valle Hermoso). **C.** TB incidence in non-border jurisdictions [Ciudad (Cd.) Victoria, Tampico, Cd. Mante, San Fernando, Jaumave, Padilla, and Altamira}.

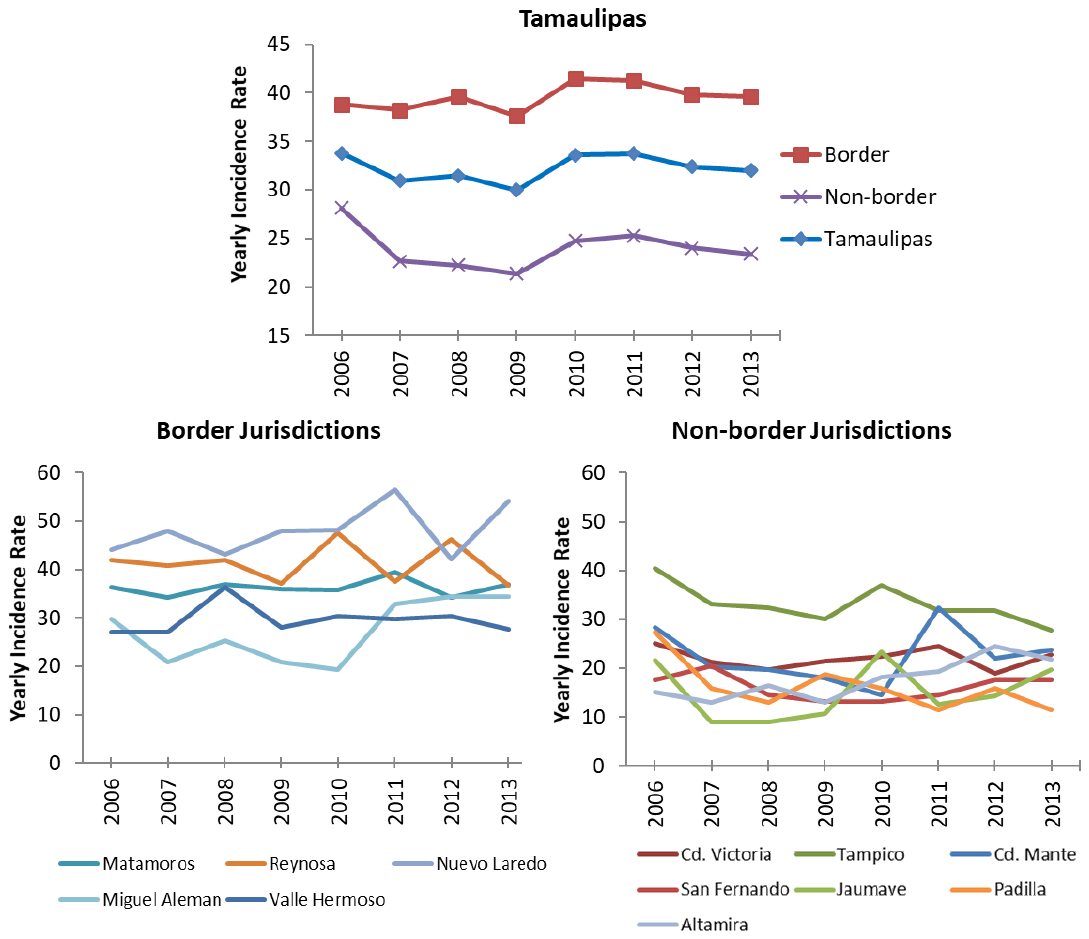
